# Performance of Wearable Pulse Oximetry During Controlled Hypoxia Induction

**DOI:** 10.1101/2024.07.16.24310506

**Authors:** Yihang Jiang, Connor Spies, Will Ke Wang, Ali R. Roghanizad, Satasuk Joy Bhosai, Laurie Snyder, Ashley Burke, David MacLeod, Jessilyn Dunn

## Abstract

Oxygen saturation is a crucial metric used for monitoring patients with lung disease or respiratory illness who are at risk of hypoxemia (low blood oxygen saturation). Early and accurate identification of abnormal oxygen saturation is important for these patients who may develop significant desaturation and hypoxemia symptoms during their daily activities. To investigate the performance of two pulse oximeters during desaturation, we recruited nine healthy participants and performed a controlled oxygen desaturation study that reduces the blood oxygen saturation levels from 100% to 60% using a gas delivery system. We calculated the oxygen desaturation rate (ODR) of each measurement and conducted a comparative analysis of the displayed oxygen saturation readings from both the Masimo MightySat Rx finger pulse oximeter and Apple Watch Series 7 with arterial blood oxygen saturation readings obtained from a blood gas analyzer. Both the Masimo MightySat Rx pulse oximeter and the Apple Watch Series 7 tended to overestimate oxygen saturation. The pulse oximeter readings were more likely to fall within 2% of the acceptable (as specified by Masimo) peripheral oxygen saturation (SpO_2_) error range than the Apple Watch (49.03% compared to 32.14%). Notably, both devices had limitations under low oxygen saturation levels (<88%) with an accuracy root mean square difference (A_rms_) of 3.52 [3.18-3.86]% and 5.82 [5.32-6.31]% for the Masimo MightySat Rx and Apple Watch Series 7, respectively. Among the blood oxygen measurements taken during a high oxygen desaturation rate (ODR) (i.e., ≥ 2% SpO_2_ per minute), which is a rate clinically correlated with sleep apnea, the A_rms_ increased slightly by 0.75% for the Masimo MightySat Rx and decreased by 0.28% for the Apple Watch Series 7. However, no statistically significant relationship was found between the oxygen desaturation rate and the device measurement errors.

## Introduction

Blood oxygen saturation, representing how much hemoglobin is bound with oxygen, is considered to be a useful vital sign for gaining insight into a person’s health status alongside body temperature, blood pressure, heart rate, and respiratory rate(1). Chronic lung diseases such as chronic obstructive pulmonary disease(COPD) and asthma can lead to significant desaturation and hypoxemia (2,3), and exercise-induced desaturation is common in these populations and associated with increased mortality (4,5). Early and accurate identification of abnormal oxygen saturation (6) is important in patients who may develop hypoxemia symptoms and significant oxygen desaturation during exercise, rest, or sleep (7–9).

Measurement of oxygen saturation can be classified into two methods: invasive and non-invasive (10). Arterial blood gas (ABG) analysis is an invasive measurement method that requires the collection of arterial blood samples and which measures arterial oxygen saturation (SaO_2_) by a dedicated co-oximeter machine. While ABG analysis is considered the ‘gold standard’, arterial blood sample collection is painful and impractical for continuous monitoring (11,12). Pulse oximetry is based on the principle that the optical absorption of light across specific wavelengths differs for oxyhemoglobin and deoxyhemoglobin and this phenomenon allows for the non-invasive estimation of blood oxygen saturation (SpO_2_) (13). SpO_2_ measurement functionality was first introduced in popular consumer wearables in 2021, and roughly a decade later in 2032, the global market value of wearable technology is projected to reach $191.58 billion (14). Multiple studies comparing SpO_2_ measurements from the Apple Watch Series 7 and finger-placed pulse oximeters found Pearson correlation coefficients ranging from 0.76 to 0.89(15–18). However, these studies did not evaluate the smartwatch’s capability to detect and identify hypoxia specifically, which is when accuracy is most critical because incorrect measurements can put patients at risk of delayed recognition and treatment(19–21). Occult hypoxemia, which occurs when a person’s blood oxygen level appears normal when measured by pulse oximetry (SpO > 92%), but is actually low when measured by a more accurate test like arterial blood gas (SaO <88%), may be a clinically significant consequence of such unreliability (22). In such a situation, hypoxemia is present but "hidden" or not detected by the pulse oximeter. Occult hypoxemia is concerning because it can delay the recognition and treatment of low oxygen levels, especially in critically ill patients or those with respiratory diseases like COVID-19.

Additionally, while SpO_2_ is often straightforward to measure when its level has reached a steady state, challenges arise in trying to accurately measure SpO_2_ under dynamic change, and particularly when those changes are rapid such as during sleep apnea (23,24). Blood oxygen levels typically decrease in people with obstructive sleep apnea (OSA) as a result of gaps in their breathing (25). The oxygen desaturation rate (ODR), defined as the change in SpO_2_ per minute, is a metric used to quantify such changes in SpO_2_ over time. During rapid desaturation, pulse oximeter accuracy has been shown to decrease due to the averaging-window algorithms that smooth the signal over several seconds and include values before the desaturation event(26,27), causing the reported SpO_2_ value to lag behind the patient’s real-time oxygenation status and resulting in incorrect values. Because pulse oximeter accuracy may be mediated by the ODR, it is important to evaluate pulse oximeter performance under conditions of rapid oxygen desaturation. While such studies have been performed in the past, they typically do not use the gold standard SaO_2_ method to measure blood oxygen saturation due to its burdensome nature of sample collection (28–30). In this study, we evaluated whether the accuracies of SpO_2_ measurements from a common pulse oximeter and a smartwatch were impacted by the ODR using gold standard SaO_2_ measurements.

## Materials and Methods

### Ethical Considerations

This study was approved by the Duke University Health System (DUHS) Institutional Review Board (Pro00110458). Written informed consent was obtained from participants prior to their participation in the study.

### Study Population

This study was conducted as an opportunistic investigation embedded within an existing controlled hypoxia study, which involved invasive arterial catheterization and prolonged exposure to reduced oxygen levels. As a result, the number of participants was constrained by the requirements of the parent study. Inclusion criteria included healthy male or female subjects between the ages of 18 and 45 years with a minimum body weight of 40 kg and a body mass index between 18.0 and 35.0 kg/m². Individuals were excluded if they had peripheral vascular disease, Raynaud’s syndrome, cryoglobulinemia, or any collagen vascular disease affecting the fingers; a history of blood clots within the past six months; a history of sickle cell trait or thalassemia; a positive urine cotinine test; heparin allergy; essential tremor; gel nail polish or any other non-natural, non-removable discoloration of the forefinger; and, based on venous blood sampling, abnormal hemoglobin levels, abnormal hemoglobin electrophoresis, carboxyhemoglobin levels greater than 3 g/dL, or methemoglobin levels greater than 2 g/dL.

### Equipment

Equipment was used in this study to assess skin tone, control the desaturation process, and take the SpO_2_ and arterial oxygen saturation (SaO_2_) measurements. To assess skin tone, the Delfin SkinColorCatch colorimeter was used to report the Individual Typology Angle (ITA) which classifies skin color types into six groups, from Very Light to Dark skin: Very Light (>55°), Light (41° to <55°), Intermediate (28° to <41°), Tan (10° to <28°), Brown (-30° to <10°) and Dark (<-30°). In the desaturation process, we used the sequential gas delivery system RespirAct RA-MR to control the alveolar oxygen tension (PAO_2_) and partial pressure of end-tidal oxygen (PetO_2_) by changing inhaled oxygen and carbon dioxide levels (31). The attained PAO_2_ determined the partial pressure of oxygen in arterial blood (PaO_2_) at the alveolar-arterial interspace in the lung. The PaO_2_ in turn determined the participants’ SaO_2_ prior to blood gas sampling. The Radiometer ABL90 Flex blood gas analyzer was used to analyze extracted arterial blood gas samples and obtain the functional arterial oxygen saturation. The Apple Watch Series 7 smartwatch (reflectance pulse oximeter) and Masimo MightySat Rx finger pulse oximeter (transmissive pulse oximeter) were each used to measure SpO_2_ separately. The Apple Watch Series 7 was chosen as it was previously identified to have the best performance among a selection of four commercial wearables in an analytic validation study (15,32,33), and the Masimo MightySat Rx was chosen because it is a FDA 510(k) cleared pulse oximeter (34,35).

### Desaturation Protocol

In this study, we employed a controlled gas delivery system to conduct the controlled desaturation protocol and altered the SaO_2_ by adjusting end-tidal oxygen (PetO_2_) levels that participants inhaled (Figure 1). We then compared the oxygen saturation (SpO_2_) measurements from Apple Watch Series 7 and the Masimo MightySat Rx against the gold standard ABG measurements.

**Figure 1.**
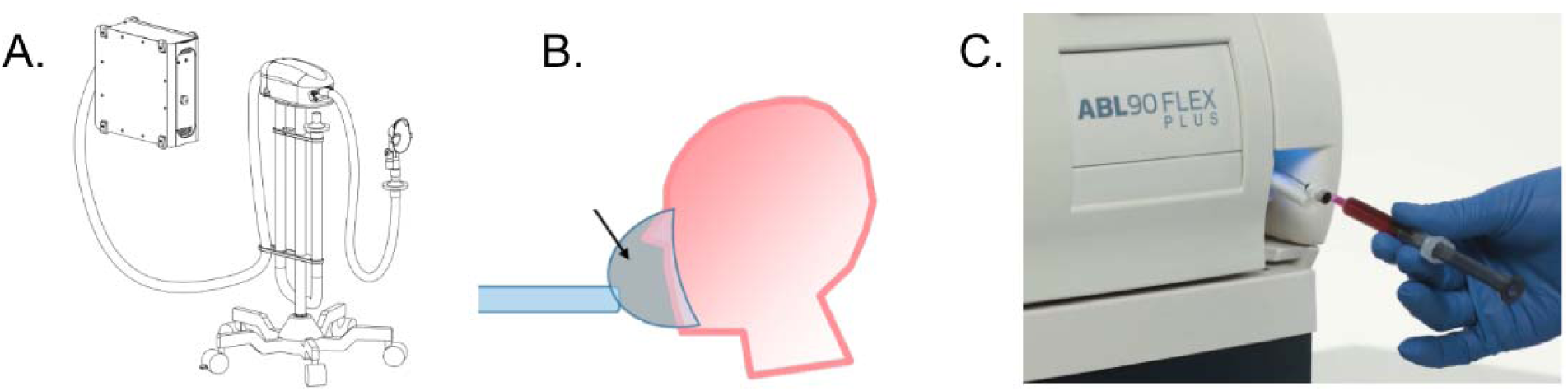
Workflow of the equipment used to adjust PetO_2_ and to measure SaO_2_. Adjusting the end-tidal PetO_2_ setting in the RespirAct RA-MR gas delivery system (A) led the study participant to inhale gas with different oxygen, carbon dioxide and nitrogen concentrations through a fitted mask (B). Blood samples taken once the PetO_2_ stabilized were processed by a blood gas analyzer (Radiometer ABL90 Flex) (C) to measure the resulting SaO_2_ and PaO_2_.

Before the desaturation procedure, participants laid in the supine position on a standard hospital stretcher with an arterial catheter placed in the forearm for blood sampling. The facemask that was connected to the sequential gas delivery system donned by the participant. The Apple Watch Series 7 was placed comfortably but tight enough on the right wrist, approximately one centimeter above the ulnar styloid process, to avoid movement during the experiment. The Masimo MightySat Rx was placed on the middle finger of the participant’s right hand.

This study involved three phases: a sequential, stepwise oxygen desaturation phase with monotonically decreasing oxygen saturation (Figure 2A, Phase 1), an oxygen saturation recovery phase (Figure 2A, Phase 2), and an interrupted oxygen desaturation phase which included a brief increase in blood oxygen saturation during the overall desaturation sequence which enables evaluation of pulse oximeters during directional changes in blood oxygen saturation (e.g., desaturation to resaturation or resaturation to desaturation) (Figure 2A, Phase 3). Besides the oxygen saturation recovery phase, Phase 1 was designed to follow a standard step-down sequence (34), as recommended by FDA, for assessing the performance of pulse oximeters during desaturation and hypoxemia while Phase 3 was designed to evaluate device performance when the desaturation was interrupted under hypoxemia. Blood oxygen levels (SaO_2_) were altered by controlling inhaled oxygen and carbon dioxide levels using the RespirAct gas delivery system, which correspondingly altered PaO_2_ and PetO_2_, in turn, SaO_2_ (36). In the first sequence, the oxygen saturation (SaO_2_) was reduced in stepwise fashion from ∼100% to ∼60%, and then back to ∼100% over the course of approximately 24 minutes. The second sequence involved holding participants’ SaO_2_ at ∼100% for approximately 30 minutes. The third sequence involved decreasing participants’ SaO_2_ from ∼100% to ∼80%, then back to ∼100% over the course of 21 minutes. The first oxygen desaturation sequence (Figure 2A, Phase 1) involved 8 steps of approximately 3 minutes each with PetO_2_ set to 90, 60, 50, 45, 40, 37, 34, 32, and 250 mmHg. Using the Severinghaus formula (37), SaO_2_ levels at these stages were estimated to be 97%, 90%, 85%, 81%, 75%, 70%, 65%, 60% and 100%, respectively. Following this sequence, PetO_2_ was held at 250 mmHg for approximately 30 minutes to allow participants to recover their maximum oxygen saturation (Figure 2A, Phase 2). The final oxygen desaturation sequence (Figure 2A, Phase 3) consisted of 7 steps of approximately 3 minutes each with PetO_2_ set to 90, 60, 50, 60, 50, 45, 40 and 250 mmHg, corresponding to an estimated SaO_2_ of 97%, 90%, 85%, 90%, 85%, 80%, 75% and 100%. During each step of the sequence, PetO was held at a steady state using the RespirAct RA-MR until two ABG samples were obtained: one collected once PetO had stabilized, and another collected one minute later, right before transitioning to the next target PetO level. Consecutive plateaus were separated by approximately two minutes to allow the gas delivery system to reach the next target PetO level. The only exception was the recovery period between Phase 2 and Phase 3, during which participants were given a 30-minute rest interval. A participant who completes the full desaturation protocol is expected to have 36 ABG samples corresponding to two samples at each of the 18 steady-state plateaus, which themselves comprised eight plateaus during Phase 1 (initial desaturation), two plateaus during Phase 2 (recovery at high PetO ), and eight plateaus during Phase 3 (interrupted desaturation).

**Figure 2.**
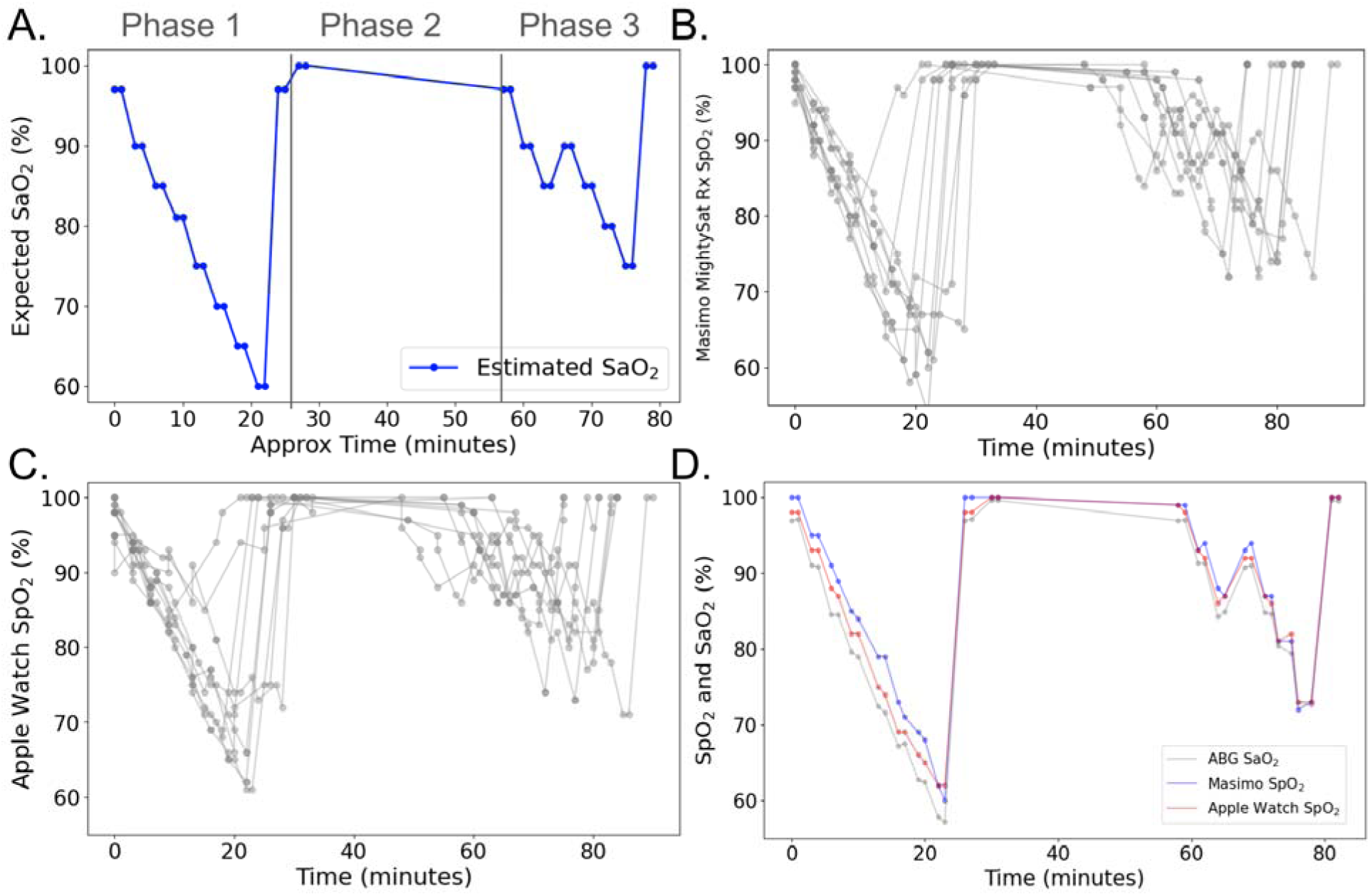
**(A)** Severinghaus-estimated SaO_2_ values. The solid blue dots represent the estimated SaO_2_ based on the PetO_2_ settings at each step of the protocol, and the solid lines show the overall trajectories. The three phases of the study protocol (1: oxygen desaturation; 2: oxygen saturation recovery; 3: interrupted oxygen desaturation) are separated using gray vertical lines. **(B and C)** Actual SpO_2_ measurements from the Masimo MightySat Rx (B) and the Apple Watch Series 7 (C) throughout the study protocol (N=9 study participants). (**D**) Actual SpO and SaO measurements from a representative participant resulting from the changing PetO in the inhaled gas mixture over the duration of the study.

Two measurements of SpO_2_, one each from the Apple Watch Series 7 and the Masimo MightySat Rx, were obtained simultaneously with each ABG sample at each steady-state plateau. Measurements on the smartwatch were manually triggered and the participants were instructed to keep their arm still during the 15-second countdown while awaiting the SpO_2_ reading. The finger pulse oximeter reading was recorded once the smartwatch produced either a successful or unsuccessful measurement. If Masimo MightySat Rx or Apple Watch Series 7 failed to produce a measurement, we recorded a missing (invalid) measurement and made another attempt after readjusting the device. If both devices successfully produced readings, the smartwatch was manually triggered again to initiate the second measurement.

### Metrics and statistical analysis

To assess the performance of the MightySat Rx and Apple Watch Series 7, we compared each SpO_2_ measurement to the SaO_2_ measurements, determining if they fell within, above, or below the acceptable error range (2%) as defined by the Masimo MightySat Rx manual. We also recorded whether the SpO_2_ measurements were missing, indicating that the pulse oximetry device was unable to generate a measurement. The Bland-Altman method was employed to assess the accuracy of SpO_2_ measurements across all readings, with separate analysis for readings where the SaO_2_ value was below 88%. The threshold of 88% was chosen in line with the British Thoracic Society guidelines (31) indicating the decision point for implementing intensive therapy to elevate oxygen saturation. The x-axes of the Bland-Altman plot are the mean of the SaO_2_ and SpO_2_ values and the y-axes are the difference between SaO_2_ and SpO_2_. Metrics to gauge each device’s performance included: Mean directional error (MDE) (Eq 1), missingness (Eq 2) and average root mean square difference (A_rms_) (Eq 3) where p is the number of participants and v_i_ is the number of valid measurements for the i^th^ participant. To evaluate device performance during oxygen desaturation, we excluded measurements with SpO_2_ or SaO_2_ >99% (n=57) as well as measurements collected during the resaturation process (n=79), which was defined by an increase in SaO relative to the preceding measurement (i.e., between Phase 1 and Phase 2 of the protocol). For the remaining measurements (n=172) with ODR ≥ 2%/min or <2%/min, we utilized a two-sided paired t-test to examine the hypothesis that MDE differs between the two ODR scenarios. The choice of 2%/min stems from the 4% oxygen desaturation index (ODI-4%), which is defined as the number of events per hour with a more than 4% decrease in SpO_2_ within two minutes (i.e., the equivalent of an average of a two percent change per minute)(24). ODI-4% is a clinically important metric to help diagnose obstructive sleep apnea (38). However, because the sampling frequency of the SaO_2_ measurements is less than one measurement every two minutes, we thus translated this threshold into an ODR of 2%/min. Before our statistical testing, the differences between each pair of SpO and SaO point measurements (see supplemental Table 1) were mean-aggregated for each person at each of the two ODRs, resulting in one MDE per person per ODR scenario. This was done separately for the Apple Watch Series 7 and for the Masimo MightySat Rx SpO measurements. These calculations resulted in nine participant-level MDE values for each ODR scenario and for each device (see Supplementary Table 1, column 7; Supplementary Figure 1; see Supplementary Figure 2 for normality check). Two paired t-tests were performed separately, one for each devic, on the nine MDE values when ODR≥ 2%/min and the nine MDE values when ODR < 2%/min to determine whether MDE values were statistically significantly different when ODR was above or below 2%/min.

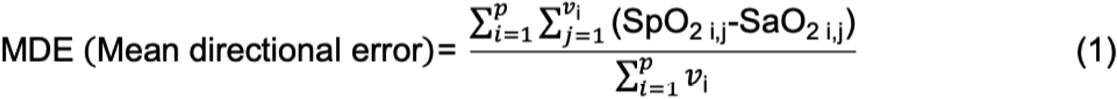

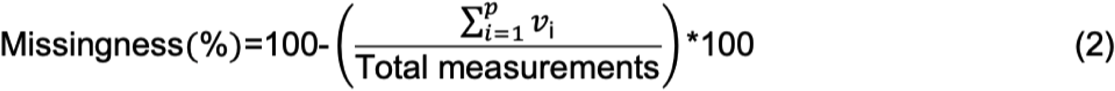

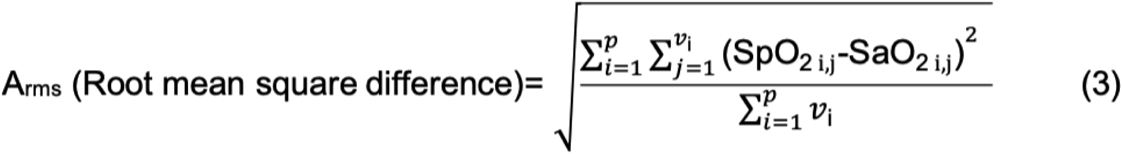

## Results

### Study Population

Nine individuals, five males and four females, provided written informed consent following the Duke University Health System IRB protocol (Pro00110458) and completed the study as described in the Methods. The ITA values of all participants ranged from -27° to 54°, with a median value of 7° (Table 1).

**Table 1.** Participant demographics and characteristics.

| Participant Number | ITA value | Reference ITA skin classification (39,40) |
| --- | --- | --- |
| 1 | 23 | Tan |
| 2 | 54 | Light |
| 3 | -27 | Brown |
| 4 | -8 | Brown |
| 5 | 12 | Tan |
| 6 | 39 | Intermediate |
| 7 | 2 | Brown |
| 8 | -1 | Brown |
| 9 | 7 | Brown |

### Comparison between Arterial Blood Gas and Pulse Oximetry Measurements

Measurements were taken across 9 study participants at 18 set PetO_2_ levels ranging from 32 to 250 mmHg, which translates to an approximate SaO_2_ from 60% to 100% over the course of three phases of the study that involved monotonic oxygen desaturation, recovery, and interrupted oxygen desaturation (Figure 2A; a representative participant’s trajectory is shown in Figure 2D). There were 308 measurements of the “ground truth” ABG-based blood oxygen saturation (SaO_2_) which were compared with the SpO_2_ measurements generated by the Masimo MightySat Rx (Figure 2B) and the Apple Watch Series 7 (Figure 2C). All participants had maximum ABG-based SaO_2_ measurements of 99% and maximum Masimo- and Apple watch-based SpO_2_ measurements of 100% (Table 2). The minimum SaO_2_ was less than 65% for 6 out of 9 (67%) participants, and minimum SpO_2_ was less than 65% for 7 out of 9 (78%) participants using the Masimo MightySat Rx and for 2 out of 9 (22%) participants using the Apple Watch Series 7. In general, the Apple Watch Series 7 measurements did not reach the same nadir values as th Masimo MightySat Rx measurements (Figure 2B and 2C).

**Table 2.**
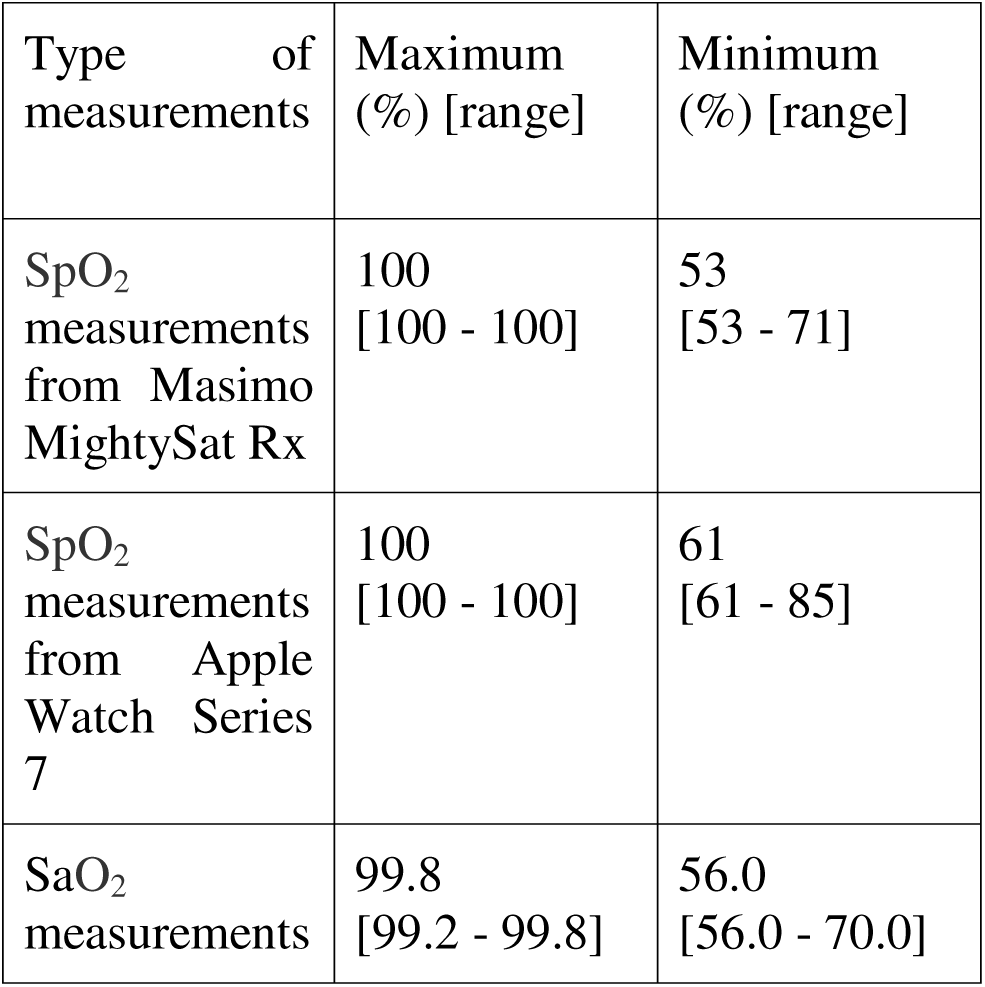
The Maximum and Minimum SpO_2_ and SaO_2_ Values and their interindividual ranges during the Desaturation Procedure.

Each measurement from the Masimo MightySat Rx and the Apple Watch Series 7 was compared to its concomitant ABG measurement to determine whether it fell within, above, or below 2% error, which was the acceptable range as defined by the Masimo MightySat Rx manual. The percentage of measurements falling into these different categories demonstrates whether there exists a trend of over- or underestimation of SpO_2_ levels. Both the MightySat Rx and the Apple Watch Series 7 tended to overestimate SpO_2_ with mean directional errors (MDE) of 1.80% and 3.26%, respectively (Figure 4, Figure 4A, Figure 4B). The Apple Watch Series 7 had a higher proportion of overestimated measurements (56.49% of SpO_2_ measurements were overestimated) as compared with the MightySat Rx (where 44.48% of measurements were overestimated). A higher percentage of Masimo MightySat Rx SpO_2_ measurements (49.03%) fell within the 2% error range of the reference SaO_2_ values compared to Apple Watch Series 7 SpO_2_ measurements (32.14%).

Arms across 5% SaO_2_ bins demonstrated a decreasing trend (Figure 3) for both devices with increasing SaO_2._ The highest Arms values were observed at the lowest saturation ranges (60–65%), reaching 7.26% for the Apple Watch Series 7 and 4.57% for the Masimo MightySat Rx. In contrast, the lowest Arms values occurred at the highest saturation range (95–100%), where Arms decreased to 2.27% for the Apple Watch Series 7 and 1.53% for the Masimo MightySat Rx. Overall, the Masimo MightySat Rx exhibited lower Arms values than the Apple Watch Series.

**Figure 3.**
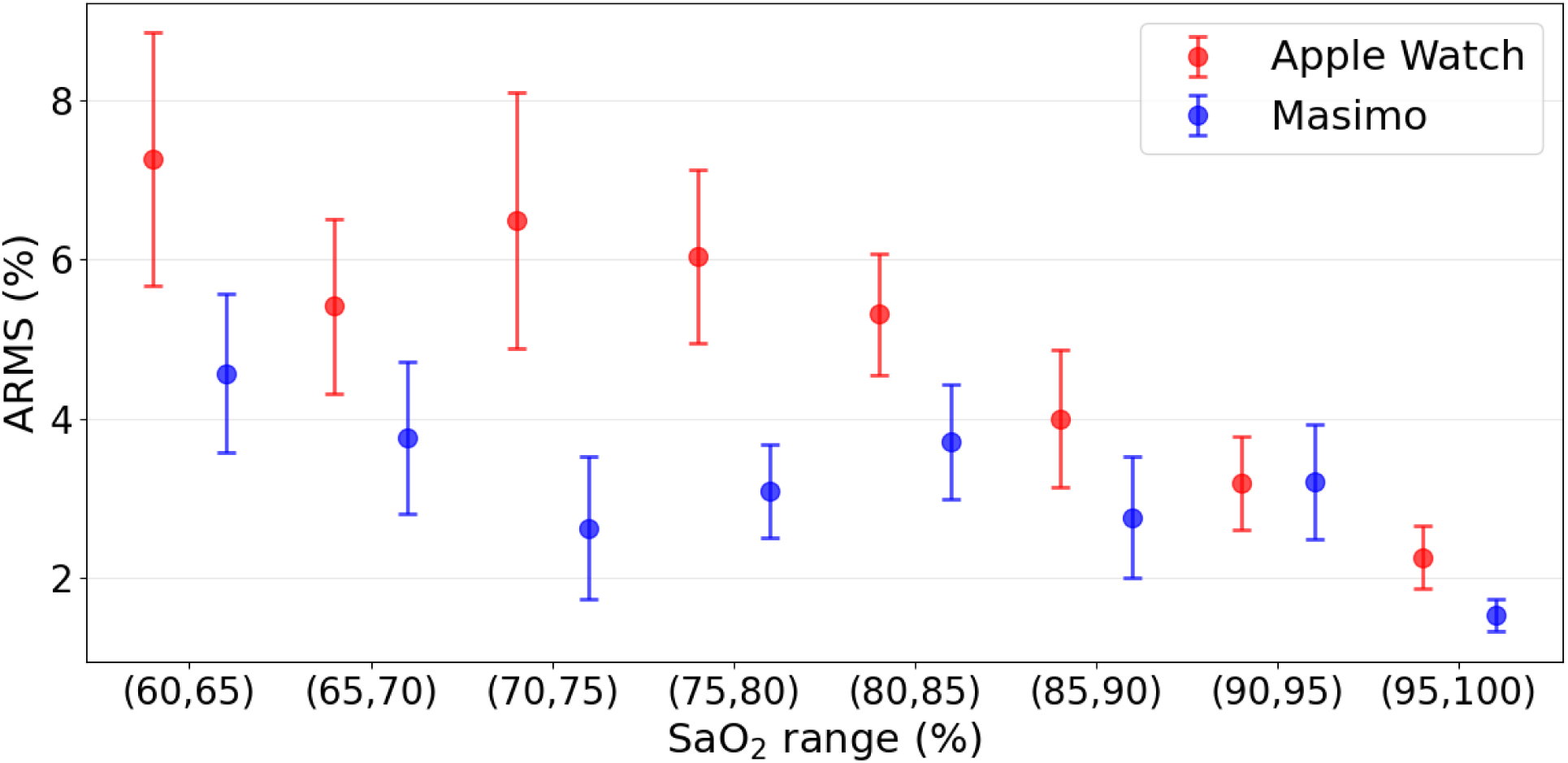
A_rms_ of SpO_2_ measurements for Apple Watch Series 7 (red) and Masimo MightySat Rx (blue) binned by 5% oxygen saturation bins ranging from 60% to 100%. Points indicate the A_rms_ within each SaO range, and error bars represent the 95% confidence intervals.

A Bland–Altman analysis demonstrated that SpO_2_ measurements tended to be overestimated since the points in the figure mostly lie above the zero-bias line (Figure 4A, Figure 4B). Overall, the Masimo MightySat Rx showed higher accuracy against the SaO_2_ ground truth measures than the Apple Watch Series 7, with an average root mean square difference in accuracy (A_rms_) [95% CI] of 2.98 [2.72-3.25]% for the Masimo MightySat Rx and 4.63 [4.26-5.00]% for the Apple Watch Series 7.

**Figure 4.**
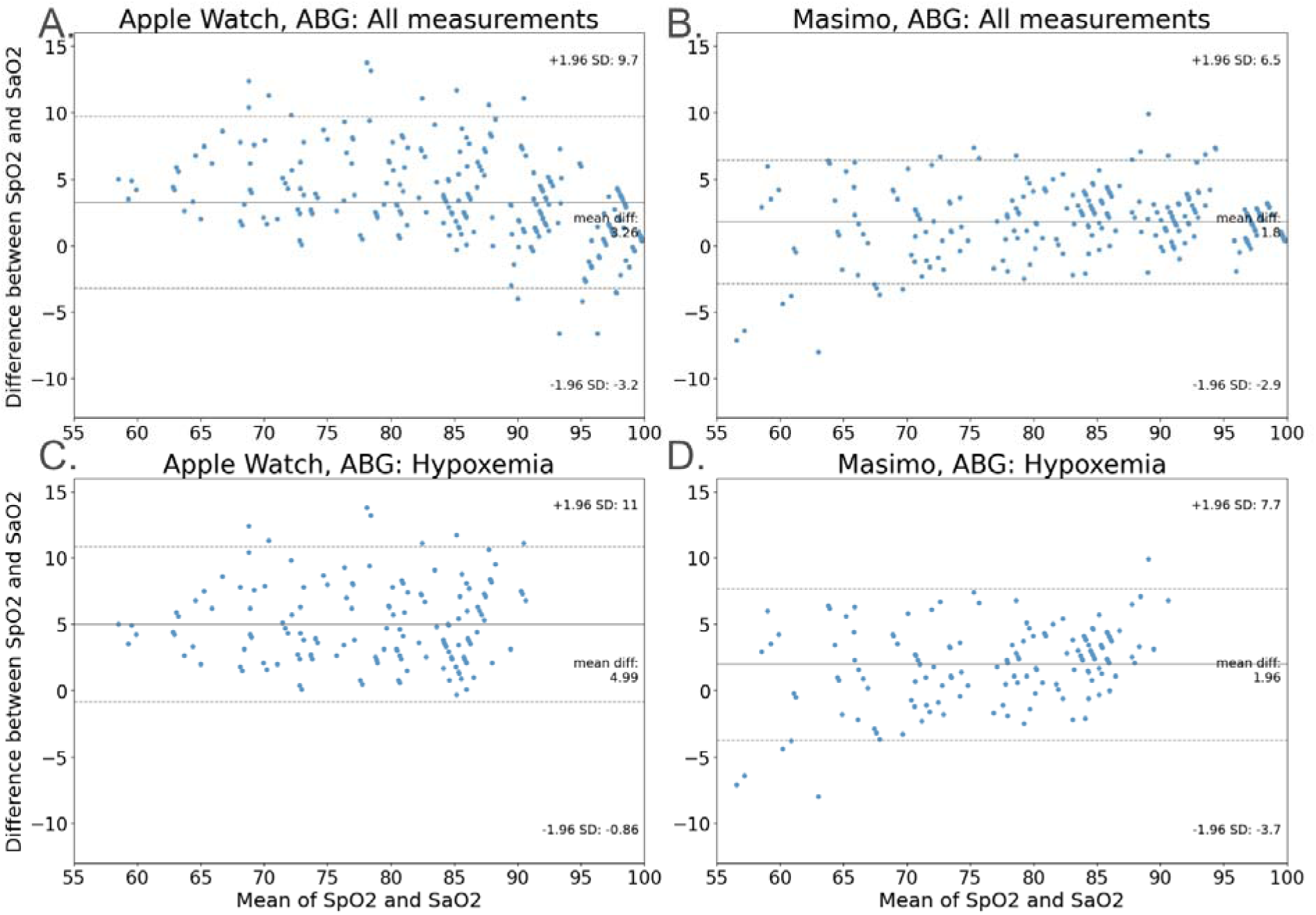
Bland-Altman plots demonstrate the differences between simultaneous ABG-based SaO_2_ measurements and either all (A) or hypoxemia-only (SaO2 < 88 %) (C) SpO2 measurement from the Apple Watch Series 7. The same comparisons are shown between SaO_2_ and SpO2 for th Masimo MightySat Rx (all, B; and hypoxemia-only, D). The solid line shows the mean difference of the measurements (MDE) and the dashed lines show the 95% limits of agreement.

### Missingness Evaluation

Of attempted SpO_2_ measurements, 305 out of 308 (99%) and 284 out of 308 (92%) were successfully reported by the Masimo MightySat Rx and Apple Watch Series 7, respectively. In other words, when the device was prompted to make a measurement, the Masimo MightySat Rx was unable to produce three out of 308 attempted measurements, while the Apple Watch Series 7 was unable to produce 24 out of 308 attempted measurements (Figure 5, blue bar). Thus, in terms of missingness, the Apple Watch Series 7 had more missingness (7.79%) compared to the MightySat Rx (0.97%), indicating that both have high likelihood (>90%) for successfully obtaining a measurement when a measurement was attempted.

**Figure 5.**
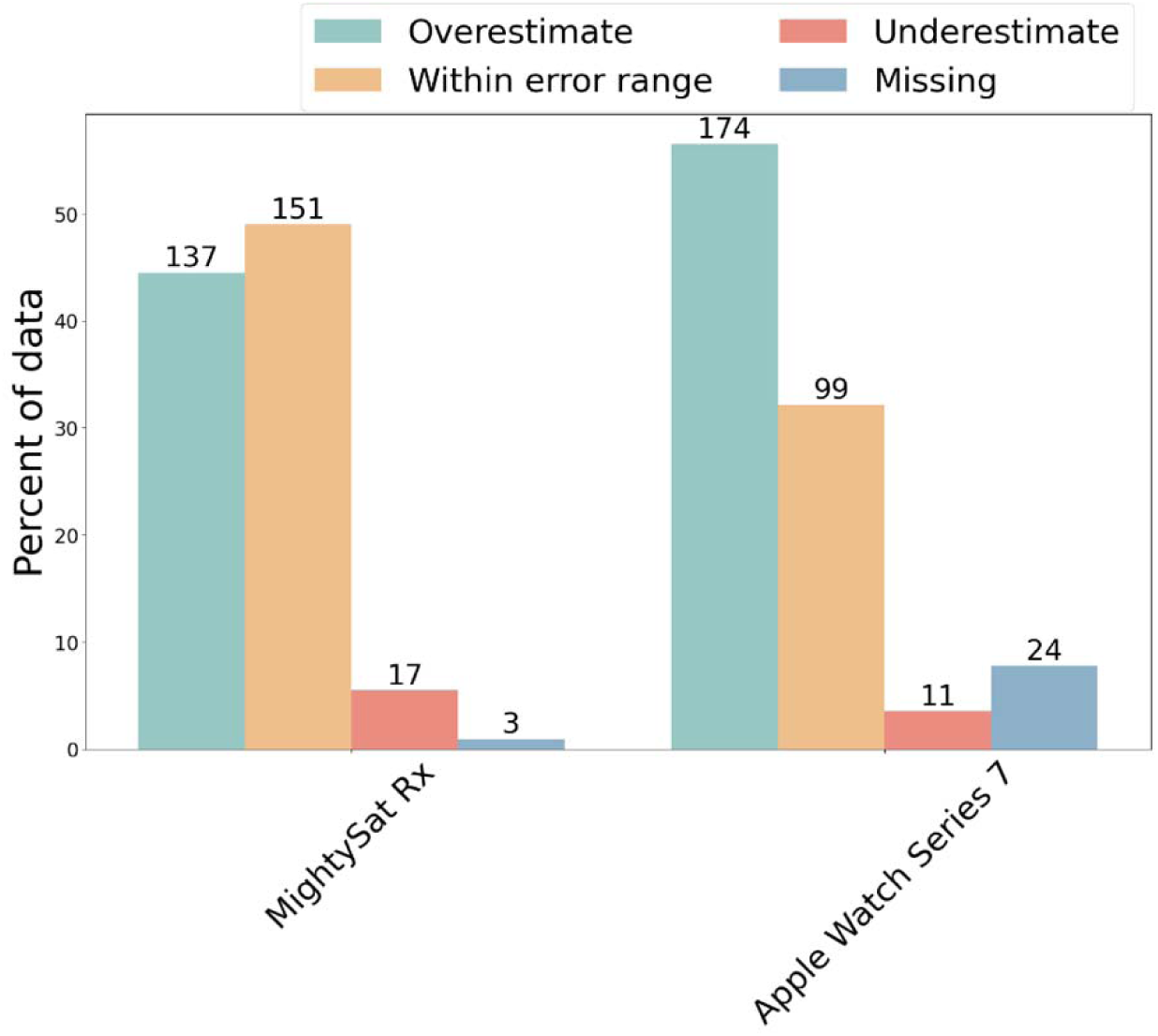
Relative percentages of data falling into the categories of overestimated measurements (green), measurements within acceptable error ranges (yellow), underestimated measurements (red), and missing measurements (blue) for the Masimo MightySat Rx and Apple Watch Series 7.

### Accuracy of measurements under hypoxemia

The clinical threshold for hypoxemia is blood oxygen saturation < 88% (41). As noted in the introduction, occult hypoxemia is a situation where a patient actually has blood oxygen saturation < 88%, but their blood oxygen saturation measurements read as higher (i.e., above the threshold that would lead to a clinical intervention). Among the 308 ABG-based SaO_2_ measurements, 165 (54%) were < 88%. Among those, the Masimo MightySat Rx and Apple Watch Series 7 produced 164 (99%) and 150 (91%) simultaneous SpO_2_ measurements, respectively– the remaining were attempted but the devices did not generate values and thus those values were considered missing. Among those SpO_2_ measurements where SaO_2_ was less than 88%, 86/164 (52.44%) of the Masimo MightySat Rx measurements and 128/150 (85.33%) of the Apple Watch Series 7 measurements were overestimated. Importantly, 3 (1.8%) of the Masimo MightySat Rx measurements and 6 (4.0%) of the Apple Watch Series 7 measurements were actually reported to be above 92%. These instances of occult hypoxemia occurred in participants 2 and 7 using the Masimo MightySat Rx, and in participants 2, 8, and 9 using the Apple Watch Series 7. Notably, only one measurement from a single participant (participant #2) demonstrated an instance of occult hypoxemia that was detected simultaneously on both devices.

Looking specifically under conditions of hypoxemia, the MDE of SpO_2_ for the Masimo MightySat Rx and the Apple Watch Series 7 was 1.96% and 4.99%, respectively– higher than the overall MDE of 1.80% and 3.26% reported above (Figure 4C, Figure 4D; Supplementary Table 1, which represents participant-level MDE of SpO_2_ for Masimo MightySat Rx and the Apple Watch Series 7). The A_rms_ [95% CI] during hypoxemia was 3.52 [3.15-3.87]% and 5.82 [5.34-6.30]% SpO_2_ for the Masimo MightySat Rx and the Apple Watch Series 7, respectively, which are also both higher than the overall A_rms_.

### Relationship between device performance and oxygen desaturation rate (ODR)

Before comparing device performance under ODR ≥ 2%/min and ODR < 2%/min conditions, we excluded 57 fully saturated measurements (SpO or SaO > 99%) and 79 resaturation measurements. Among the remaining 172 desaturating SaO measurements (Supplementary Table 1) out of the total 308 measurements, 169 and 157 SpO measurements were collected from the Masimo MightySat Rx and the Apple Watch Series 7, respectively. Among those, 83/169 Masimo MightySat Rx and 78/157 Apple Watch Series 7 measurements occurred when the ODR was greater than two percent per minute.

When the ODR increased above 2%/min, the Masimo MightySat Rx A_rms_ increased from 3.04% to 3.79% and the MDE increased from 1.86% to 2.38%. Also at ODR>2%/min, the Apple Watch Series 7 MDE increased from 4.16% to 4.41% (Supplementary Figure 2) but A_rms_ decreased from 5.50% to 5.22%. No significant correlation was found (Figure 6) between the ODR and MDE for the Masimo MightySat Rx nor the Apple Watch (two-sided paired t-test: P = 0.12 and 0.22, respectively).

**Figure 6.**
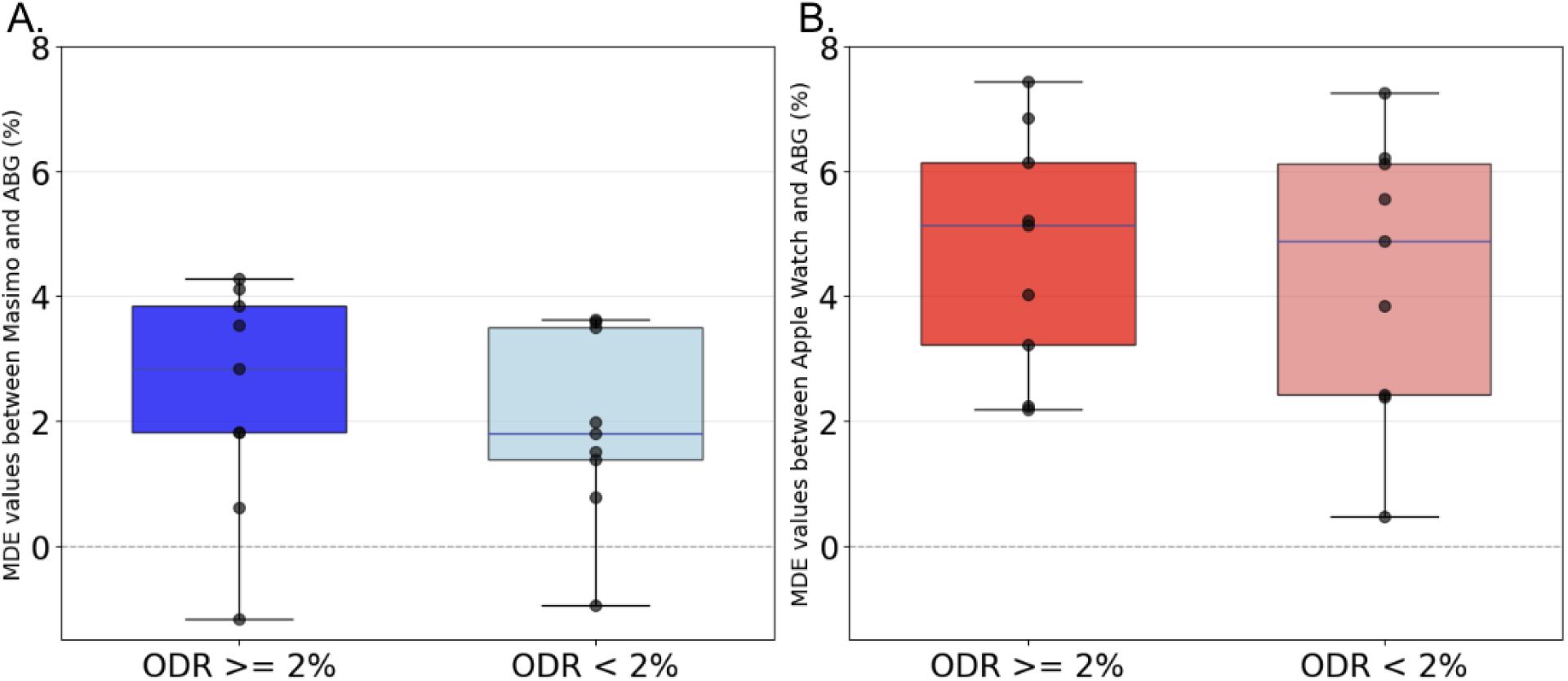
Boxplots demonstrate the participant-level MDEs of (A) Masimo MightySat Rx and (B) Apple Watch Series 7 between SpO and SaO measurements when oxygen desaturation rate ≥ 2%/min and < 2%/min.

## Discussion

We designed and implemented this study to evaluate the performance of optical blood oxygen saturation measurements (SpO_2_) from smartwatches and finger pulse oximeters under conditions of hypoxemia that were induced by a controlled oxygen desaturation protocol. There are three main findings of our study: (1) Both the Masimo MightySat Rx and the Apple Watch Series 7 often overestimate true blood oxygen saturation values, with 44% and 56% of their measurements in this study, respectively, found to be higher than the ground truth SaO_2_ measures; (2) Both devices have lower accuracy during hypoxemia than at normal blood oxygen saturation; (3) Both devices showed a trend toward lower accuracy when the ODR is larger than 2%/minute but the difference is not statistically significant.

Our previous study (32), which used the Masimo MightySat Rx as the reference measurement for oxygen saturation to analytically validate four commercial wearables in 49 patients with low blood oxygen saturation, many of whom had chronic pulmonary disease, found that the Apple Watch Series 7 overestimated blood oxygen saturation measurements 17% of the time. In this current study, where we used ABG measurements of SaO_2_ as the reference standard “ground truth” measure, as well as a controlled oxygen desaturation study protocol in healthy individuals to achieve low blood oxygen saturation, the Apple Watch Series 7 overestimated blood oxygen saturation 56% of the time. In this study, we also observed that the Masimo MightySat Rx, the reference standard used in the previous study, also overestimated oxygen saturation levels 44% of the time. One likely reason for the discrepancy in the Apple Watch Series 7 overestimation rate between the two studies is the choice of the reference standard measurement – the ABG measurements used in the current study are a more trustworthy ground truth measurement. Another likely factor is the difference in the distribution of oxygen saturation values between the two studies. Specifically, our previous study did not include a desaturation protocol, and the SpO measurements obtained from the Masimo MightySat Rx ranged only from 82% to 100%, which may have contributed to a lower observed rate of overestimation. Blood oxygen saturation overestimation is particularly dangerous under conditions of occult hypoxemia (where the actual SaO_2_ is < 88% but the measured SpO_2_ is > 92%), when a patient’s blood oxygen saturation is thought to be normal and care is delayed (42). In our study, although the Apple Watch Series 7 had a high rate of overestimation (56% of measurements), only 6/150 (4.0%) of the Apple Watch Series 7 measurements and 3/164 (1.8%) of the Masimo MightySat Rx measurements actually fell into the category of occult hypoxemia. It is likely that, had there been more measurements concentrated only slightly below 88%, there would have been more occurrences of occult hypoxemia observed in our study. This is because the mean bias of the Apple Watch Series 7 measurements under hypoxemia conditions (4.99%) just slightly exceeds the minimum difference (4%) between SpO_2_ and SaO_2_ that defines occult hypoxemia (92% vs 88%).

Our study also demonstrates that the performance of both the finger pulse oximeter and the smartwatch suffers during hypoxemia as compared to conditions of normal blood oxygen saturation. The overestimation rate increased from 44.48% to 52.44% for the Masimo MightySat Rx and from 56.49% to 85.33% for the Apple Watch Series 7 while the A_rms_ increased from 2.98% to 3.52% and 4.63% to 5.82% during hypoxemia for the two devices, respectively. For 510(k) clearance, the FDA currently recommends an A_rms_ of <3.0% for transmission pulse oximeters (e.g., Masimo MightySat Rx) and <3.5% for reflectance oximeters (e.g., Apple Watch) (34). According to the FDA’s 2025 draft guidance, the A_rms_ specification for both sensor types will be standardized to <3.0% (43). Both devices used in our study exhibited A_rms_ values exceeding the FDA threshold under conditions of hypoxemia. A previous study that recruited 50 healthy adults and conducted oxygen desaturation cycles from approximately 100% to 70% SaO evaluated the accuracy of Apple Watch SpO measurements using ABG as the gold standard; this study also reported a trend of overestimation in the Apple Watch Series 6, with an Arms of 2.18% (95% CI: 1.55–2.84%), meeting current or upcoming FDA Arms requirements. The missingness in their study was 5.29% (964 of 1020 attempted measurements gave readings) as compared with 7.79% (284 of 308 attempted measurements gave readings) in our study, likely due to more measurements in our study attempted at lower oxygen saturation (44). Another study involving 167 participants with acute exacerbation of COPD reported an MDE of 0.46% between Apple Watch Series 6 measurements and ABG-based SaO_2_ measurements (30).

When the ODR exceeded 2%/min, A_rms_ increased from 3.04% to 3.79% for the Masimo MightySat Rx and decreased from 5.50% to 5.22% for the Apple Watch Series 7. Even though the difference between SaO_2_ and SpO_2_ was not found to be statistically significant during desaturation, the fact that A_rms_ exceeded 3% for both devices indicates that they should be used with caution, particularly under rapid desaturation conditions. Accurate performance during oxygen desaturation is crucial not only for monitoring oxygen levels but also for clinical decision making. This finding is especially relevant for home monitoring in patients with obstructive sleep apnea, where the number of desaturation episodes (i.e., a drop in mean oxygen saturation of ≥ 4% over the course of 120 seconds) is linked to apnea and hypopnea (24). Inaccuracies and missingness during desaturation could result in misclassification of these episodes, leading to inappropriate assessment and treatment plans for patients.

Although current FDA guidance recommends validation of pulse oximeter accuracy within the SpO range of 70% to 100%, our results highlight the importance of evaluating device performance at lower oxygen saturation levels. Device accuracy declined substantially as SaO decreased from 100% to 60%, with Arms increasing from 2.27% to 7.26% for the Apple Watch Series 7 and from 1.53% to 4.57% for the Masimo MightySat Rx. These results suggest that limiting validation to SpO values ≥70% may overlook performance limitations in more severe hypoxemia, where accurate measurement is critical for timely clinical interventions. Expanding regulatory validation requirements to include lower saturation ranges may therefore provide a more comprehensive assessment of pulse oximeter performance.

### Limitations

The main limitation of this study is the small sample size, which resulted in insufficient representation across skin tone categories and age ranges. According to the FDA guidance for pulse oximeters, 10 or more healthy participants that vary in age and gender should be included and a minimum of 2 subjects, or 15% of the study participant pool, need to be darkly pigmented (34). However, in our study, only four (light, intermediate, tan, brown) out of the seven possible ITA skin types (very light, light, intermediate, tan, brown, dark and very dark) were represented. A benefit of our study, however, was the use of objective skin tone measures using a device rather than the more common subjective human assessment using a comparison scale like Fitzpatrick or Monk (Supplementary Figure 4). Moreover, only the brown and tan skin types were represented by more than one participant, and we did not have any representation from the dark, very dark, and very light categories. This lack of diversity across skin tones limits the generalizability of our findings to individuals with lighter or darker skin tones. We recognize this as a limitation in our study as a result of the limited time and resources available to perform the study more comprehensively.

While our study captured a wide range of oxygen saturation values from 60% to 100%, covering the FDA-recommended SpO accuracy validation range of 70% to 100%, we did not measure the response time of the finger pulse oximeter, and the Apple Watch Series 7 reported SpO_2_ readings after a fixed 15-second countdown. Previous studies have shown that finger pulse oximeters may have a delay in displaying the blood oxygen saturation values during the onset of hypoxia (45). Several factors can contribute to this delay, including the measurement site and poor peripheral perfusion (45). Other research suggests that fingertip oximeters have the slowest response in desaturation changes from normoxemia to hypoxemia (46). Future studies could consider evaluating the response time during desaturation events to ensure that pulse oximeters provide not only accurate but also timely measurements.

Although our experiment simulated the process of oxygen desaturation in healthy participants, there are additional factors to consider that may prevent these results from generalizing to patients of older age and/or who may experience oxygen desaturation as a result of a health condition. The age range of our study participants was between 19 and 28 years, COPD is most prevalent in people aged 65–84 (47). Arterial stiffness of older people results in changes in the propagation of the pulse to the periphery, thereby influencing the peripheral pulse timing and shape characteristics which can affect the performance of pulse oximeters (48).

Finally, this protocol cannot demonstrate SpO_2_ measurement performance over long periods since each participant only spent 60 minutes in the first oxygen desaturation sequence and 48 minutes in the second one. While it would be infeasible to induce oxygen desaturation for such a long period in healthy adults, clinically, it is necessary to be able to consistently monitor oxygen saturation values overnight, for example in COPD patients who often experience nocturnal hypoxemia (2,49). It is therefore necessary to perform clinical validation among COPD patients with nocturnal desaturation, defined as having a SpO_2_ value below 90% for more than 30% of the time in bed during one or more nights (50,51). Future clinical validation studies should thus perform real world monitoring over longer periods, and particularly during sleep, in COPD and sleep apnea patients to capture nocturnal desaturation caused by those conditions (52).

## Conclusion

In this controlled hypoxemia study, we evaluated how well the Masimo MightySat Rx and the Apple Watch Series 7 oxygen saturation measurements (SpO ) correspond to arterial blood gas measurements (SaO ) across a range of arterial oxygen saturation levels, ranging from approximately 60% to 100%. Both devices consistently overestimated SpO, with accuracy declining notably during hypoxemia. The Apple Watch Series 7 mean bias suggests a likelihood for missing instances of hypoxemia, particularly at SaO values below, but close to, 88%. Both the Apple Watch Series 7 and Masimo MightySat Rx exhibited A_rms_ values exceeding the FDA threshold under conditions of hypoxemia. While past studies have implicated high oxygen desaturation rates in increasing measurement error, we found no statistically significant relationship between ODR and measurement error for either device. Overall, our findings of SpO overestimation and high A_rms_ values underscore the need for caution when interpreting oxygen saturation values from these devices. The small sample size and limited diversity in skin tone and age restrict the generalizability of our findings. Future studies should include larger and more diverse populations to evaluate the performance of wearable-based pulse oximetry.

## Data Availability

Deidentified data used for submission is available after acceptance.

## Acknowledgements

This study was supported in part by funds from AstraZeneca (LS and SJB), by the National Institute of Diabetes and Digestive and Kidney Diseases of the National Institutes of Health (R01DK133531 to JD), and by NSF CAREER #2339669. The content is solely the responsibility of the authors and does not necessarily represent the official views of AstraZeneca or the National Institutes of Health.

## Data Availability

The datasets analyzed during this study are available from the corresponding author upon reasonable request.

## Abbreviations

A_rms_: Average Root Mean Square
SpO_2_: Peripheral oxygen saturation
SaO_2_: Arterial oxygen saturation
PetO_2_: Partial pressure of end-tidal oxygen
PAO_2_: Alveolar oxygen tension
PaO_2_: Partial pressure of oxygen in arterial blood
ABG: Arterial blood gas
ODR: Oxygen desaturation rate

